# Interplay of ADHD polygenic liability with birth-related, somatic and psychosocial factors in ADHD - a nationwide study

**DOI:** 10.1101/2021.08.18.21262211

**Authors:** Isabell Brikell, Theresa Wimberley, Clara Albiñana, Bjarni Jóhann Vilhjálmsson, Esben Agerbo, Anders D. Børglum, Ditte Demontis, Andrew J. Schork, Sonja LaBianca, Thomas Werge, David M. Hougaard, Merete Nordentoft, Ole Mors, Preben Bo Mortensen, Liselotte Vogdrup Petersen, Søren Dalsgaard

## Abstract

**Background:** ADHD is multifactorial, yet the interplay ADHD polygenic risks scores (ADHD-PRS) and other ADHD associated risk-factors remains relatively unexplored. The aim of this study was to investigate associations, confounding and interactions of ADHD-PRS with birth, somatic and psychosocial risk-factors previously associated with ADHD.

**Methods:** Participants came from the Danish iPSYCH2012 case-cohort, including a randomly selected general population sample (N=21,578), and all ADHD cases with an ICD-10 diagnosis F90.0 (N=13,697), born in Denmark 1981-2005. We derived ADHD-PRS and identified 25 ADHD risk-factors in Danish national registers. Logistic regression was used to estimate associations of ADHD-PRS with each risk-factors in the general population. Cox models were applied in the full case-cohort to evaluate confounding of risk-factor associations by ADHD-PRS and family psychiatry history, and interactions between ADHD-PRS and each risk-factor.

**Results:** ADHD-PRS was associated with 14 out of 25 ADHD risk-factors in the general population, e.g., maternal autoimmune disorder, mild traumatic brain injury (TBI), and most psychosocial risk-factors. In the full case-cohort, 21 risk-factors were associated with ADHD diagnosis. Adjusting for ADHD-PRS and parental psychiatric history only led to minor attenuations of these associations. Interactions were observed between ADHD-PRS and sex, maternal autoimmune disease, TBI, paternal employment and age at child-birth.

**Conclusion:** Higher ADHD-PRS is associated with exposure to certain birth and somatic ADHD risk-factors, and broadly to psychosocial adversity. Evidence of gene-environment interactions were weak and ADHD-PRS and/or family psychiatric history have limited confounding effect on ADHD risk-factor associations, suggesting that majority of the investigated risk-factors act largely independently of ADHD-PRS to increase risk of ADHD.

## Introduction

Attention-deficit/hyperactivity disorder (ADHD) is a prevalent, often persistent neurodevelopmental disorder affecting 5-10% of children and 2.5-5% of adults.^1, 2^ Family, twin, and genome-wide association studies (GWAS) have demonstrated the importance of genetic factors in ADHD, with heritability estimated at 70-80% is twins and 22% from single nucleotide polymorphisms (SNP-based heritability [h^2^_SNP_]).^3, 4^ Several birth-related (e.g., low birth), somatic (e.g., infections, seizures), and psychosocial (e.g., low family income, parental psychiatric disorders) factors have also been associated with risk of ADHD.^5-8^ However, the complex interplay between ADHD polygenic liability and such risk-factors (defined here as any attribute, characteristic, or exposure of an individual that increases the likelihood of developing ADHD) is not well understood.^6^ Given that many ADHD risk-factors are themselves heritable, risk-factor associations may in part or fully be mediated by shared genetic effects influencing both the exposure and ADHD. Such effects will be captured by associations between ADHD polygenic liability (both at the family and individual level) and the risk-factor (i.e. gene-environment correlations), and by attenuated risk-factor outcome associations when accounting for ADHD polygenic liability (i.e. genetic confounding). The impact of a risk-factor may also vary by polygenic liability (i.e., gene-environment interaction). Examining these three types of gene-environment interplay is important to understand the impact of genes and specific environments on ADHD, and their joint effects, and to identify causal and potentially amenable risk-factors. ^9, 10^

ADHD is highly polygenic, meaning that hundreds or thousands of common genetic variants, each of small effect size, contribute to its etiology. Thus, polygenic risk scores (PRS), which capture the sum of an individual’s risk alleles weighted by their effect size identified through GWAS, is a potentially useful tool for investigating gene-environment interplay.^9^ A recent meta-analysis^11^ found ADHD-PRS to be associated not only with ADHD, but also with measures of lower socioeconomic status. Findings from a population-based cohort of >7000 mothers also suggest that ADHD-PRS is associated with certain prenatal risk-factors previously associated with neurodevelopment disorders (e.g. infections during pregnancy).^12^ However, research evaluating gene-environment associations is still lacking for many ADHD risk-factors, particularly in the somatic disease domain.^11^

Studies comparing differentially exposed twins and siblings have shown that many risk-factor outcome associations can be largely explained by unmeasured familial genetic and/or environmental confounding.^5^ This suggests that ADHD polygenic liability, measured at the level of the family or the individual, may influence the risk of exposure to certain ADHD risk-factors, even in the absence of an ADHD diagnosis. Parental psychiatric history is often included as a proxy of familial (genetic and/or environmental) confounding in epidemiological studies of ADHD, whereas ADHD-PRS provides a more direct proxy of ADHD polygenic liability captured by current GWAS. However, only a few studies have investigated genetic confounding in ADHD risk-factors associations using both ADHD-PRS and family psychiatric history.^13^ Similarly, there is limited research evaluating gene-environment interactions using ADHD-PRS, with exposures largely limited to socioeconomic indicators and parenting style.^11^ Recent work in the Danish iPSYCH2012 cohort, the world’s largest genotyped ADHD sample, found that both ADHD-PRS and several psychosocial risk-factors, including parental psychiatric history, were associated with increased risk of ADHD, yet there was limited evidence of interaction.^13^

In this study, we evaluate gene-environment interplay in the risk of ADHD for 25 birth-related, somatic, and psychosocial factors risk-factors associated with ADHD^5-8^ and measured in Danish national registers. First, we estimated gene-environment associations between ADHD-PRS and each risk-factors in the general population. Second, we examined the extent to which associations between the risk-factors and ADHD case-control status in iPSYCH2012 are confounded by ADHD-PRS and parental psychiatric history. Finally, we evaluated gene-environment interaction between the risk-factors and ADHD-PRS on ADHD case-control status.

## Methods and Materials

### Sample

The Integrative Psychiatric Research consortium (iPSYCH) is a case-cohort sample nested within the Danish population born between 1 May 1981 and 31 December 2005.^14^ From this study base, a 2% (N=30000) random population sample (referred to as the subcohort) and all individuals with an ADHD diagnosis F90.0 (coded according to the International Classification of Disease [ICD] 10^th^ edition) were selected. Due to the random selection, 288 individuals with ADHD were selected both as cases and in the subcohort. ADHD diagnoses was obtained from the Danish Psychiatric Central Research Register (DPCRR), which includes psychiatric hospital admissions since 1969 and outpatient admission since 1995.^15^ Genotyping in iPSYCH has been described elsewhere.^14^ For details on imputation, principal components (PC) analysis, and quality control, see Schork *et al*., (2019).^16^ iPSYCH2012 can be linked to Danish national population registers using the unique personal identification number assigned to everyone registered in Denmark. We restricted our sample to individuals alive and living in Denmark at age five, who were unrelated (no closer than third degree kinship estimated using KING v1.9) and of European ancestry, leaving 13697 ADHD individuals and 21290 non-ADHD individuals in the subcohort. A flowchart describing the study population is presented in Figure S1.

### Birth-related, somatic, and psychosocial risk-factors

To identify birth-related, somatic, and psychosocial risk-factors robustly associated with ADHD in prior literature, we conducted a targeted review of the literature including phenotypic, familial and common genetic variant studies (see Table 1 and supplementary Methods S1). From this review, we selected 25 risk-factor available in Danish national registers. The risk-factors and evidence of association with ADHD are presented in Table 1.

**Table 1.**
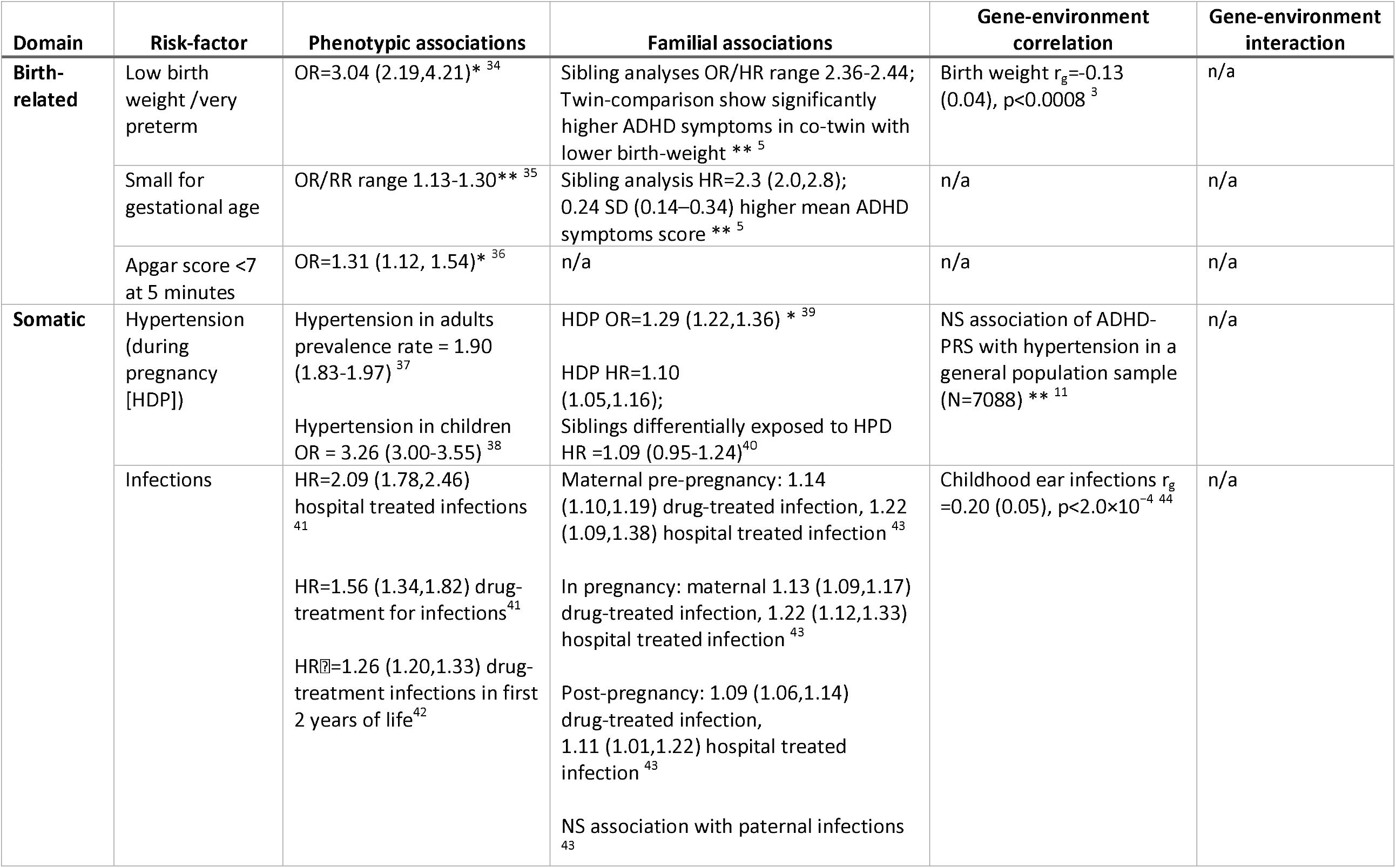
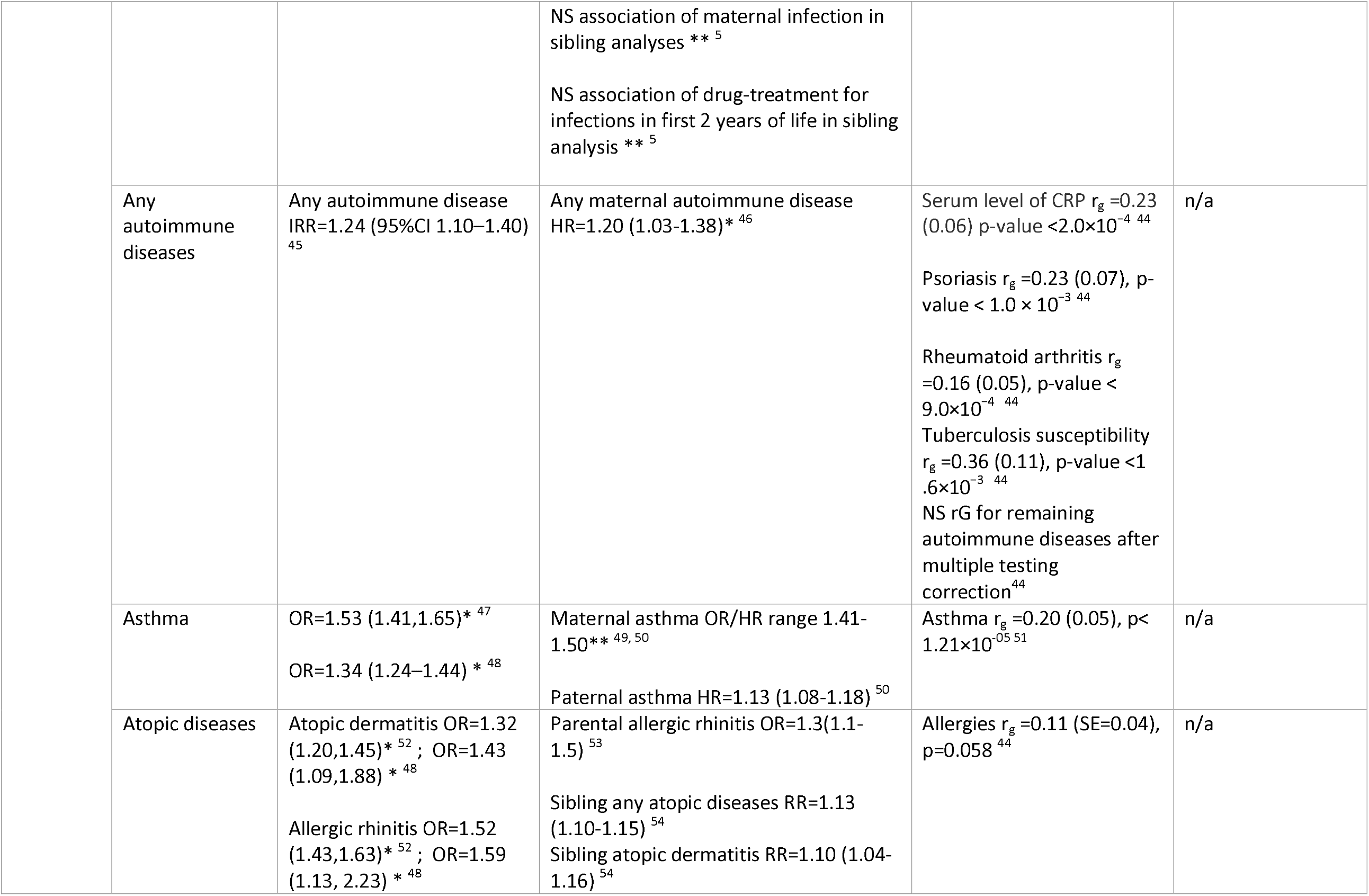
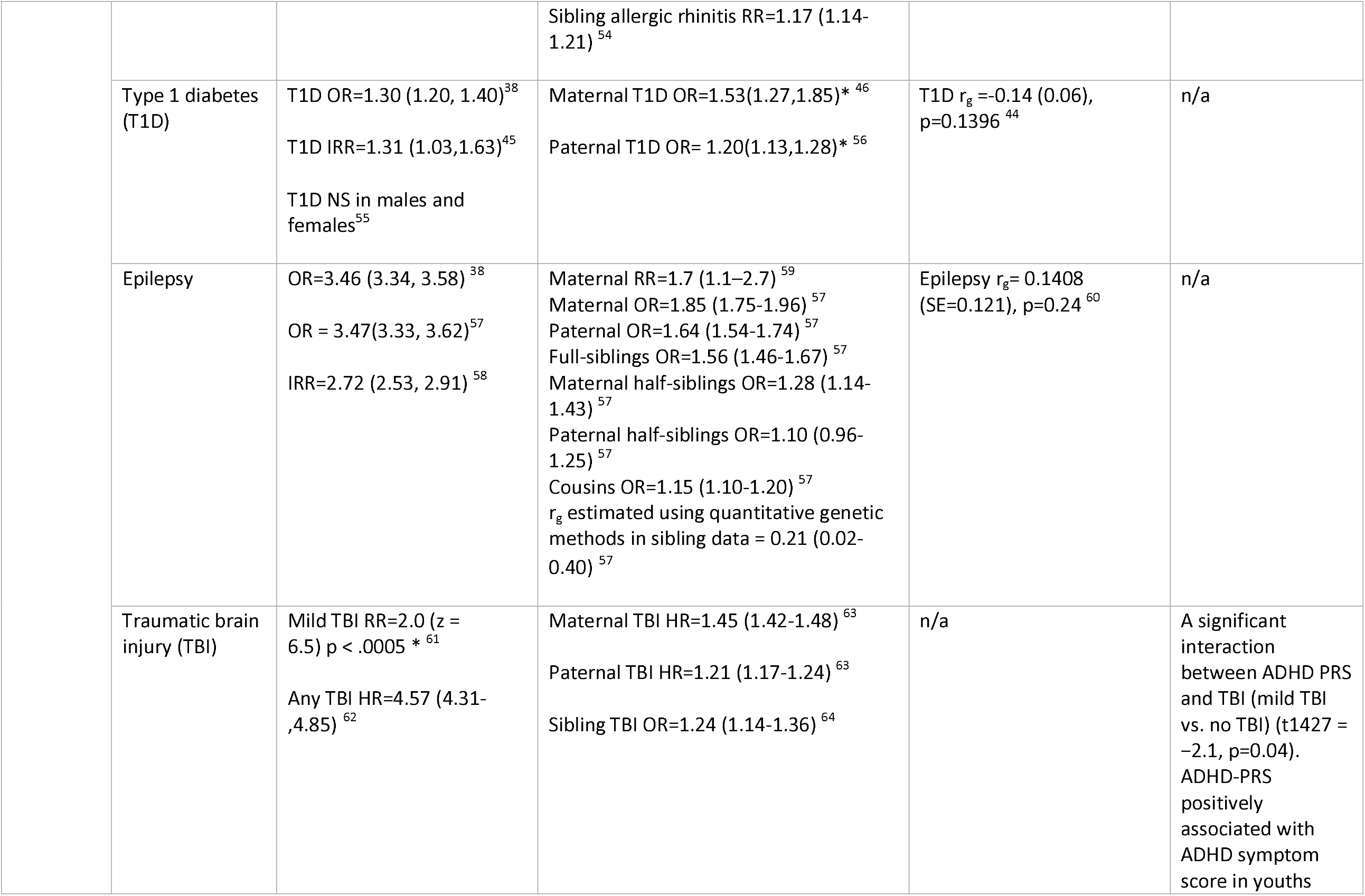
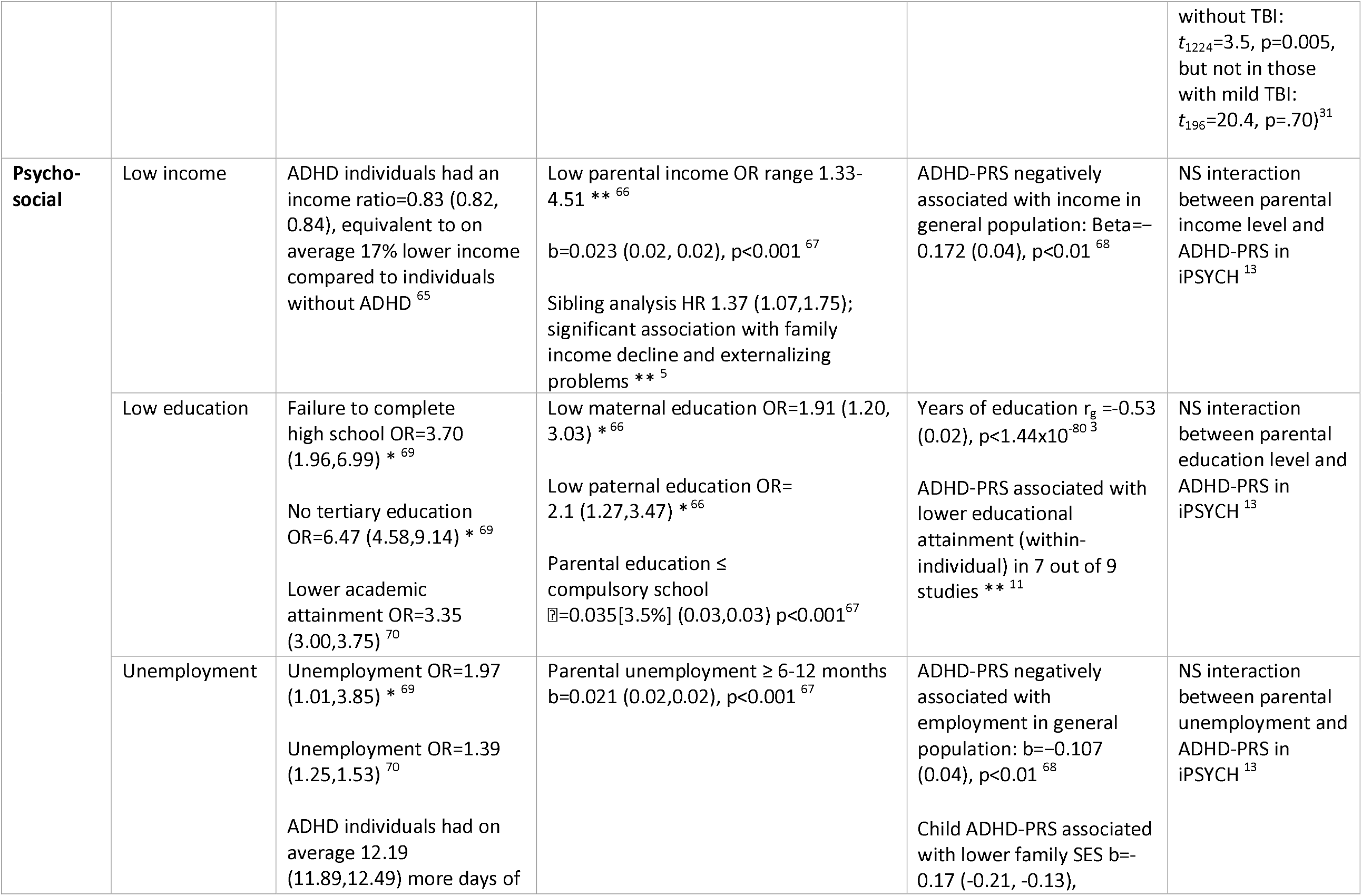
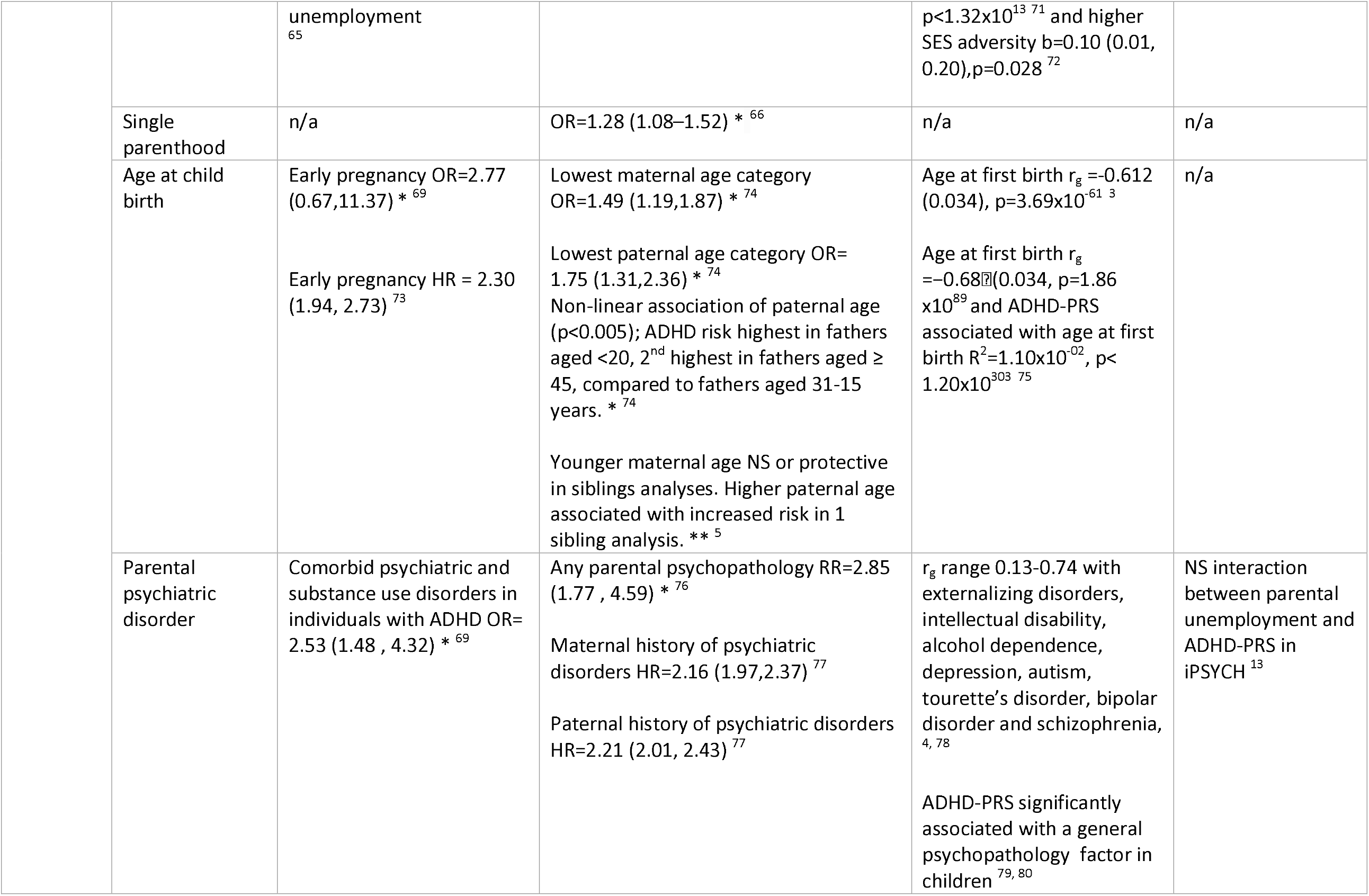
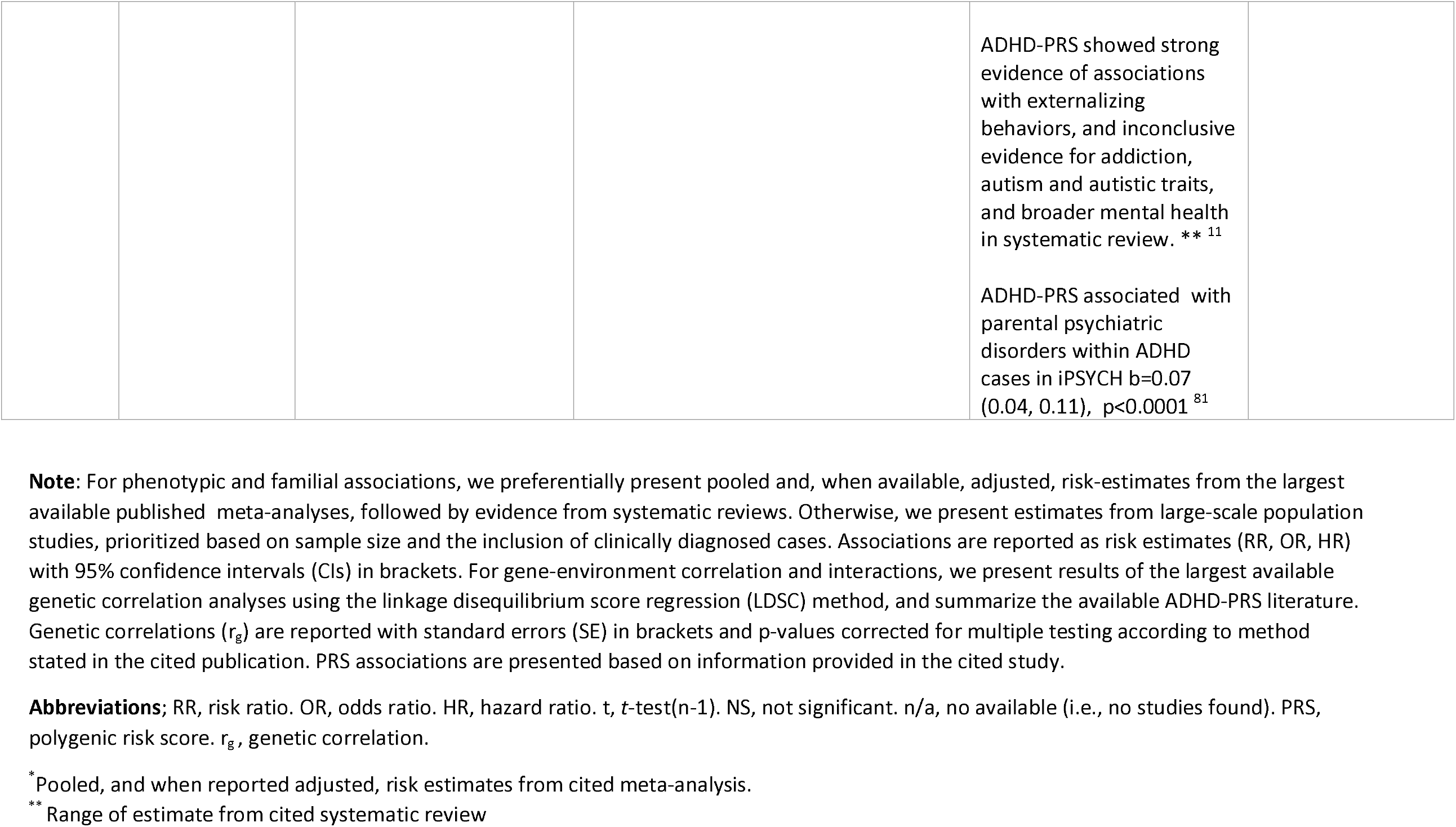
Summary of prior literature on the phenotypic, familial and genetic links between ADHD and the studied risk factors.

We extracted information on sex, date of birth, migration, death, and parents’ personal identification numbers from the Danish Civil Registration System.^17^ We used the Danish Medical Birth Register (DMBR) to obtain information on birth weight, gestational age and 5-minute Apgar score.^18^ Somatic diseases in parents (maternal hypertensive disorders and infections during pregnancy, parental autoimmune disorders) and index child (infections, asthma, atopic disease, type 1 diabetes, epilepsy, and traumatic brain injury [TBI]) were identified in The Danish National Patient Register (DNPR), which includes ICD-coded inpatient care since 1977 and outpatient since 1995.^19^ Because asthma and atopic diseases are not routinely treated in specialist care, we also used drug prescriptions from the Danish National Prescription Registry (DPR), which includes information on all prescriptions redeemed at Danish pharmacies since 1995, to identify asthma and atopic disese.^20^ Psychosocial factors (parental income, education, employment status, single parent household, age at delivery) were defined using data from Denmark’s socioeconomic registers.^21^ Finally, we parental history for any psychiatric disorder was identified using the DPCRR. Risk-factor definitions are provided in Table S1.

### ADHD polygenic risk scores

We derived ADHD-PRS in iPSYCH201 based on a combination of an externally trained PRS and an internally trained PRS. For details on the method see Albiñana et al (2021).^22^ Briefly, we used LDPred^23^ the derive an externally trained ADHD-PRS, using SNP weights from the Psychiatric Genomics Consortia cross-disorder ADHD GWAS, not including iPSYCH2012.^24^ We also leveraged having access to individual-level SNP data on a large number of ADHD individuals by calculating an internally trained ADHD-PRS in an unrelated, European ancestry subset of the iPSYCH2012 sample. SNP weights for the internally trained PRS were obtained from a mixed model prediction implemented in BOLT-LMM.^25^ The final ADHD-PRS was a linear combination of the internally and externally trained ADHD-PRS variables, standardized to the mean and standard deviation in the subcohort.

The iPSYCH2012 study was approved by the Danish Scientific Ethics Committee, the Danish Health Data Authority, the Danish Data Protection Agency, and Danish Newborn Screening Biobank Steering Committee. The Danish Scientific Ethics Committee, in accordance with Danish legislation, has, for this study, waived the need for informed consent in biomedical research based on existing biobanks.^14, 26^

### Statistical analyses

To obtain population representative estimates of the associations between ADHD-PRS and each ADHD risk-factor, we ran logistic regressions in the iPSYCH2012 subcohort, i.e. a randomly selected general population sample including 288 ADHD individuals. Associations are presented as odds ratios (OR) with 95% confidence intervals (CIs) per standard deviation (SD) of ADHD-PRS, adjusted for sex, birth-year, and for ancestry using the first four PCs. Exposures that vary over time were defined at the time of ADHD-diagnosis or end of follow-up, whichever came first.

To estimate whether associations between the risk-factors and ADHD may be confounded by ADHD-PRS and parental history of psychiatric disorders, we first confirmed their individual association with ADHD diagnosis. We then used weighted Cox models to estimate the associations of the other 24 risk-factors with ADHD case-control status, adjusted for sex, birth-year and the first four PCs (Model 1). To evaluate confounding by polygenic liability for ADHD, we further adjusted the associations for ADHD-PRS (Model 2), parental psychiatric history (Model 3), and both (Model 4). Analyses were run in the full case-control sample, i.e. all ADHD individuals (n=13697) and non-ADHD individuals from the subcohort (n=21290), with age as the underlying time-scale and Kalbfleich and Lawless weights to account for the oversampling of cases.^27^ Individuals were followed from age five until their first ADHD diagnosis, death, emigration, or end of follow-up (i.e. 31 December 2012), whichever came first. Associations are presented as incidence rate ratios (IRRs) with 95% CIs. In all Cox models, somatic and neurological risk-factors were modelled as time-varying exposures, and remaining risk-factors as time-fixed covariates defined at birth, or prior to the index child’s 5^th^ birthday.

Finally, we investigated the potential (multiplicative) interaction between each risk-factor and ADHD-PRS (modelled as a mean centered continuous variable [mean=0,SD=1]) on the risk of ADHD cases-control status, by modelling a differential linear effect of ADHD-PRS across levels of each risk-factor, using weighted Cox models, adjusted for sex, birth-year, and the first four PCs.

## Results

The full case-cohort population consisted of 34,987 individuals (14,108 females and 20,879 males), including 13697 diagnosed with ADHD (3606 females and 10,091 males). Mean age at first ADHD diagnosis was 15.6 years in females and males 13.0 years in males.

### Gene-environment associations

In the randomly selected general population sample (N=21,578, including 288 ADHD individuals), ADHD-PRS was significantly associated with 14 out of the 25 ADHD risk-factors (Figure 1 and Table S2). Among birth-related factors, higher ADHD-PRS was associated with being small for gestational age (OR=1.08, 95%CI=1.03-1.13). Among somatic risk-factors, higher ADHD-PRS was associated with maternal autoimmune disorder (OR=1.14, 95%CI=1.05-1.24), having had one (OR=1.07, 95%CI=1.05-1.10) and five or more infections (OR=1.14, 95%CI=1.05-1.24, and mild TBI (OR=1.11, 95%CI=1.05-1.17). Higher ADHD-PRS was also associated with majority of the family psychosocial risk-factors, e.g., income in the lowest quintile (maternal OR=1.19, 95%CI=1.14-1.24; paternal OR=1.19, 95%CI=1.14-1.24), low education (maternal OR=1.16, 95%CI=1.12-1.20; paternal OR=1.17, 95%CI=1.13-1.21), living in single parent household the first five years of life (OR=1.26, 95%CI=1.16-1.36), age at childbirth <20 (maternal OR=1.29, 95%CI=1.17-1.42; paternal OR=1.25, 95%CI=1.04-1.50), and parental psychiatric history (1 parent OR=1.13, 95%CI=1.07-1.20; 2 parents OR=1.30, 95%CI=1.05-1.61).

**Figure 1.**
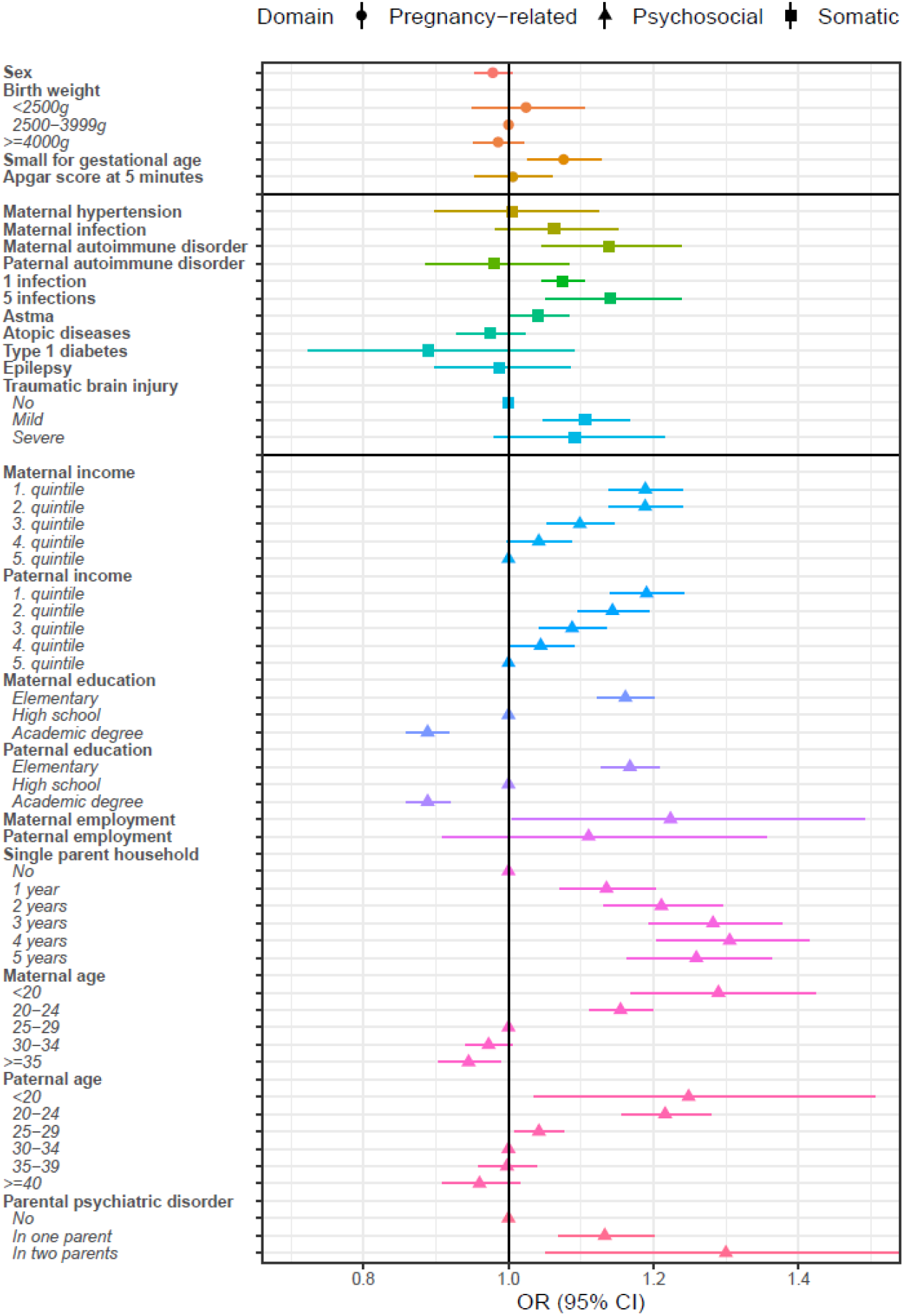
**Gene-environment associations of ADHD polygenic risk score with ADHD risk-factors in the iPSYCH2012 subcohort (N=21**,**578), expressed as odds ratios (OR) with 95% confidence intervals (CIs)** **Note**: Odds ratios and 95%CI reflect the increase in risk of exposure by one per standard deviation increase in the ADHD-PRS. Significant associations are highlighted in bold. ORs for binary exposures (0/1) are shown without reference. ORs for exposures with ≥ three levels are shown with reference. For exposure definitions, see Table S1. **Abbreviations**: OR, odds ratio. LCI, lower 95% confidence interval. UCI, upper 95% confidence interval. PRS, polygenic risk score

### Genetic confounding

In the full case-cohort, individuals in the highest ADHD-PRS decile had an increased risk of ADHD, compared to those in the lowest (IRR=4.42, 95%CI=3.96-4.93), as did individuals with two parents with a history of psychiatric disorders, compared to none (IRR=3.29, 95%CI=2.46-4.41) (Figure S2). Twenty-one risk factors were associated with an increased risk of ADHD (Table 2). Within each domain (birth-related/somatic /psychosocial) the strongest associations were observed for low birthweight (<2.5kg) (Model 1 IRR=1.85, 95%CI=1.65-2.08), epilepsy (Model 1 IRR=2.38, 95%CI=2.06-2.75), and parental at elementary school level or less (Model 1 maternal IRR=3.47,95%CI=3.24-3.72; paternal IRR=3.68, 95%CI=3.42-3.96). Maternal hypertensive and autoimmune disorders, and atopic disease and type 1 diabetes in the child were not statistically significantly associated with ADHD, with CIs including one. Adjusting the associations for ADHD-PRS (Model 2) or parental psychiatric history (Model 3) alone resulted in very minor changes. Adjusting for both ADHD-PRS and parental psychiatric history (Model 4) lead to attenuation only of the IRR’s (i.e., non-overlapping CI’s for minimally adjusted Model 1 and fully adjusted Model 4) for maternal (Model 4 IRR= 2.96, 95%CI=2.75-3.17) and paternal education at elementary school level or less (Model 4 IRR= 3.15, 95%CI=2.92-3.40), compared to an academic degree.

**Table 2.**
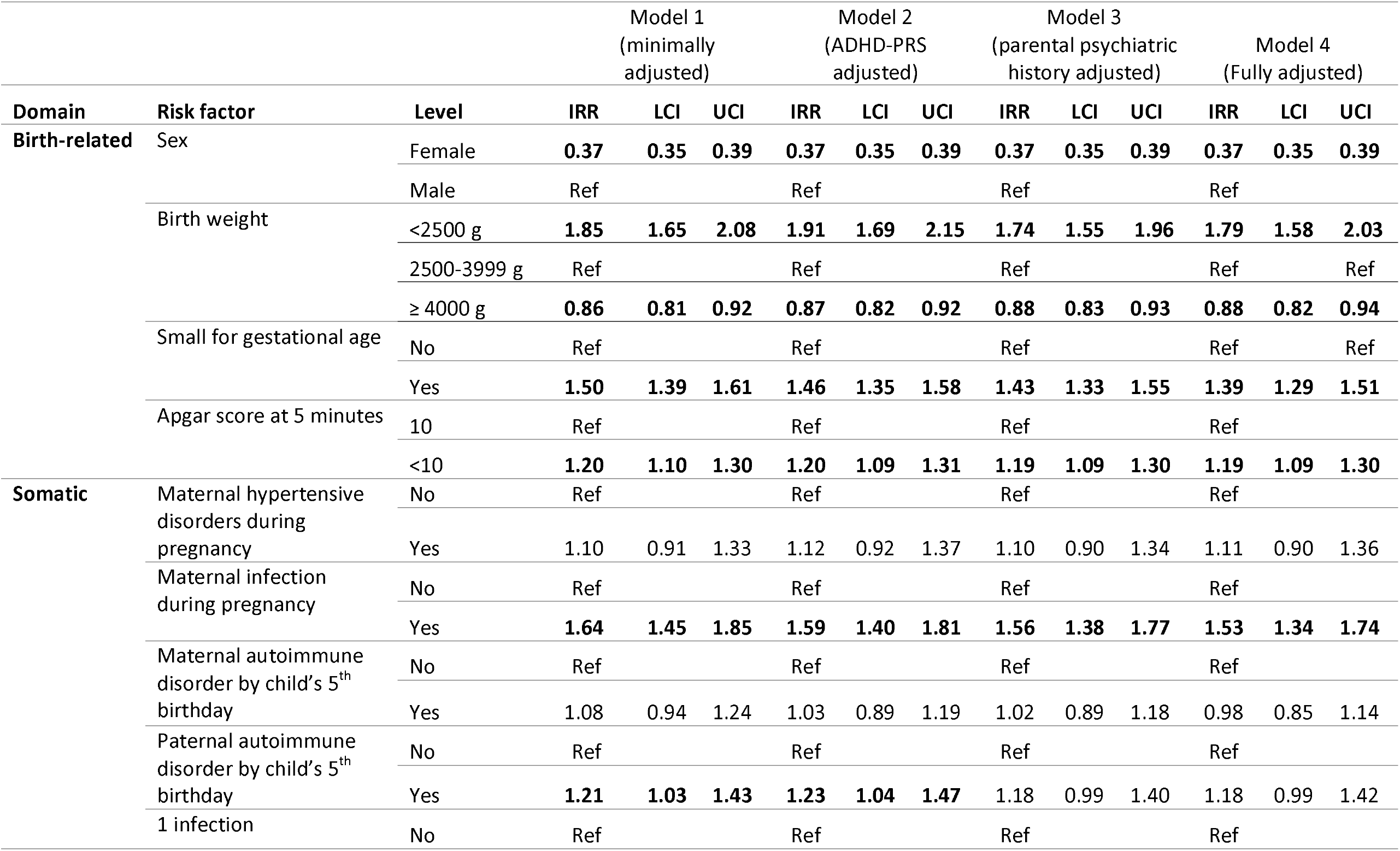
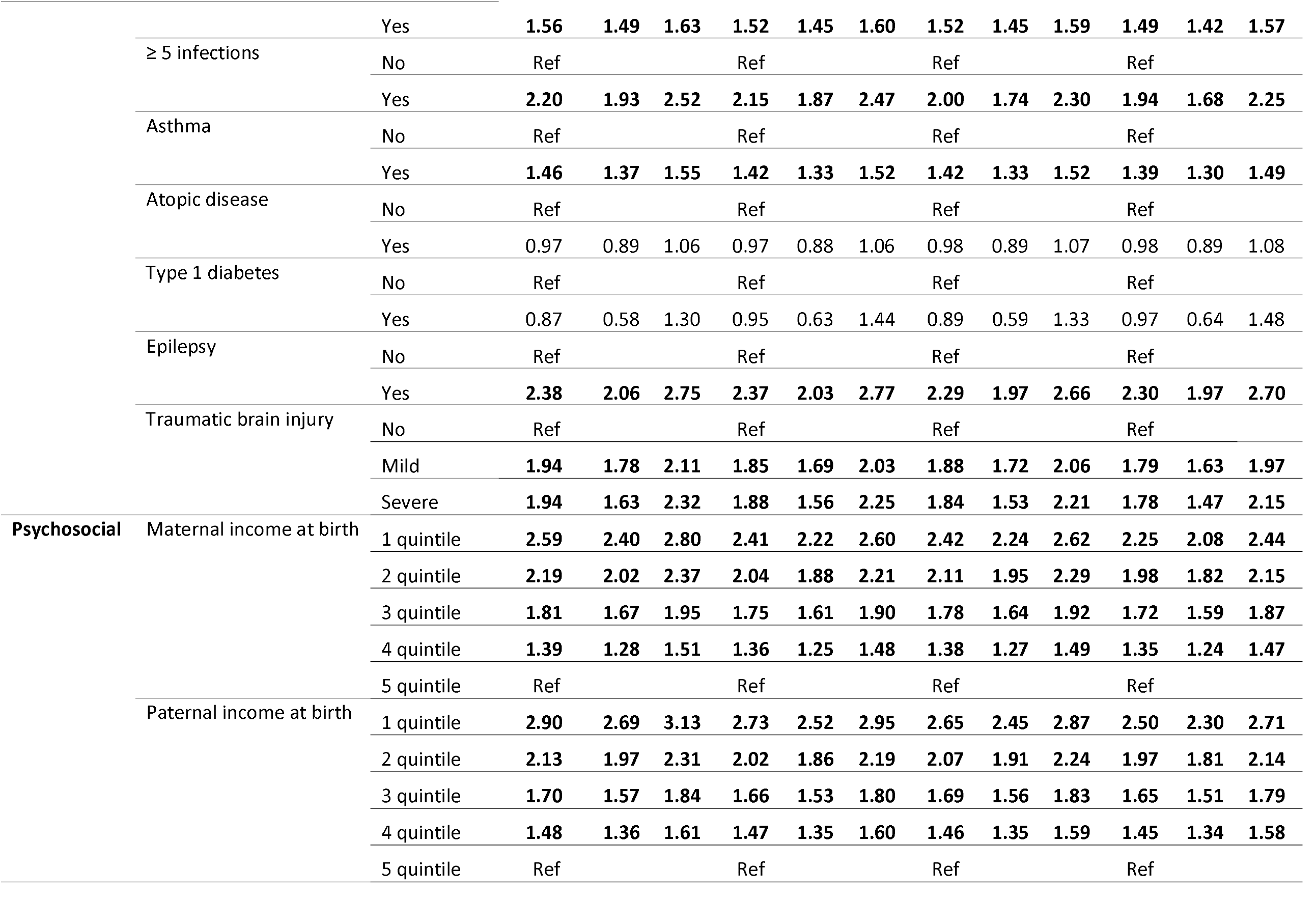
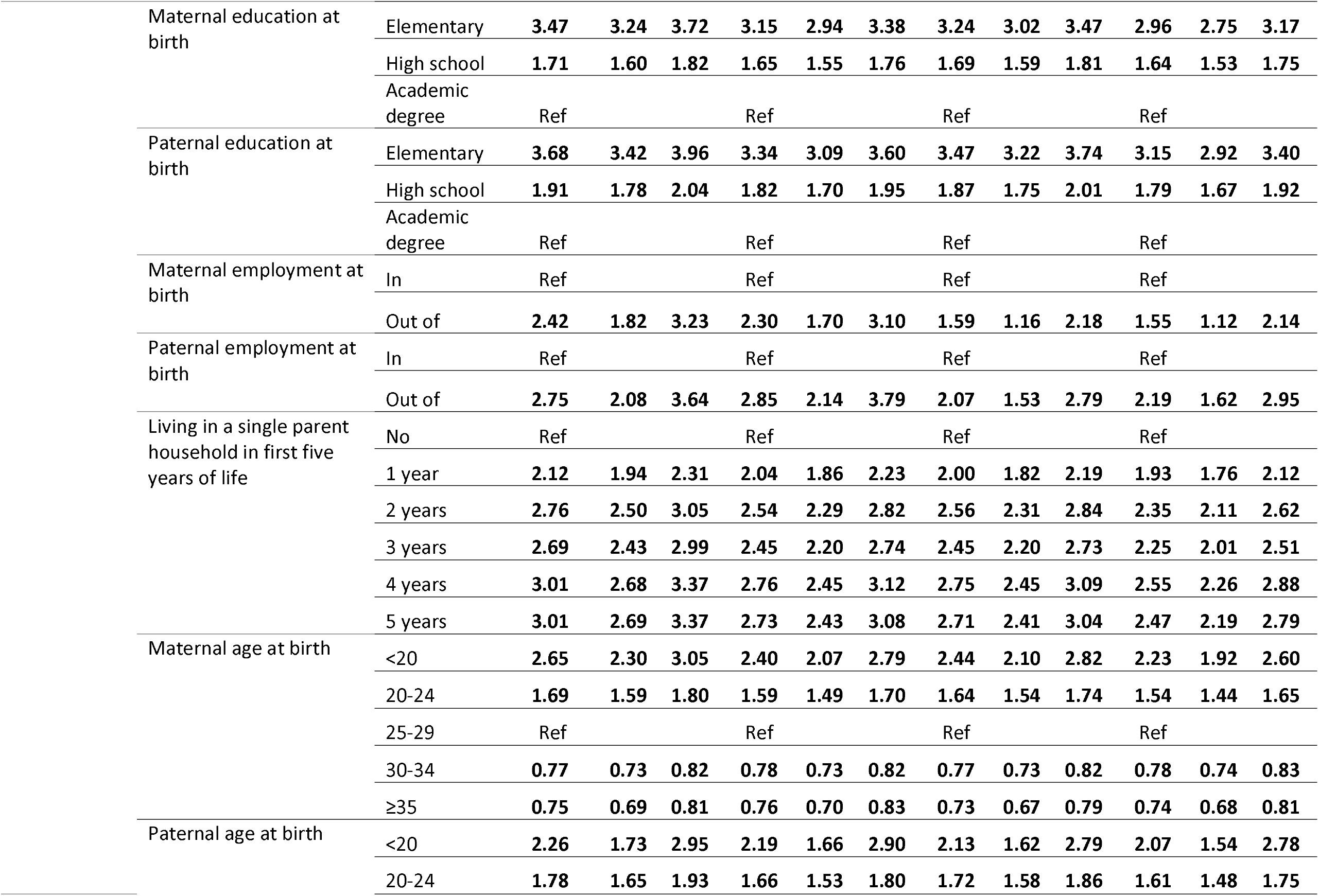
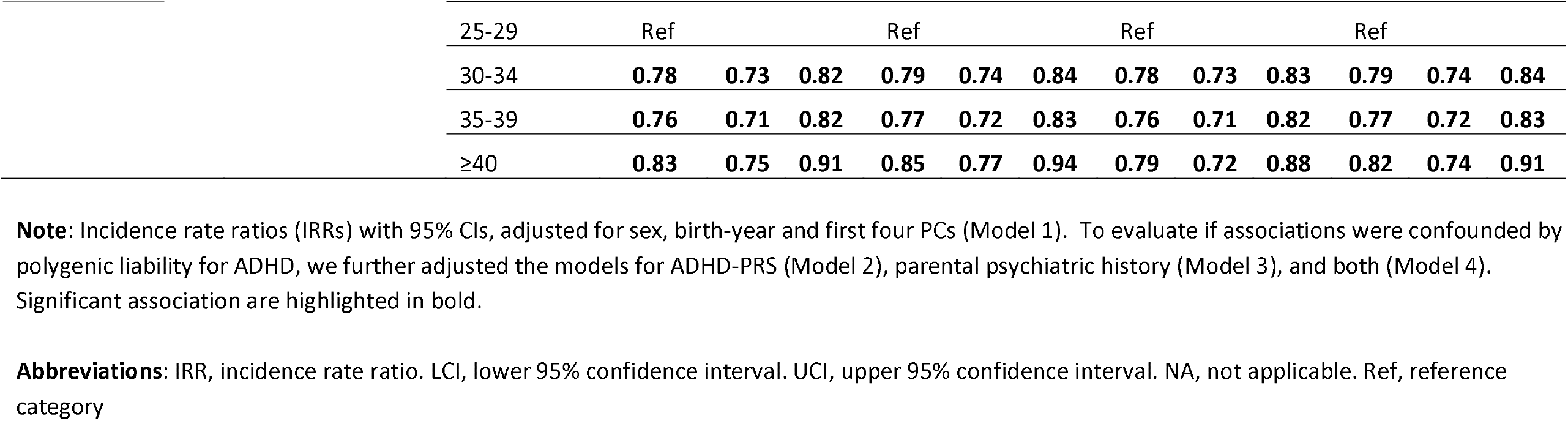
Genetic confounding of ADHD risk-factors associations with ADHD case-control status, expressed as incidence rate ratios (IRRs) with 95% confidence intervals (CIs)

### Gene-environment interaction

Results from the Cox models including interaction terms between the continuous ADHD-PRS and each risk-factor on the risk of ADHD are presented in Table S3. Evidence of interaction (*p*-value <0.05) was observed between ADHD-PRS and sex, maternal autoimmune disease, TBI, paternal unemployment and paternal age at birth of index child (Figure 2). For example, higher ADHD-PRS had a somewhat larger effect on risk of ADHD in females (Females IRR=1.60, 95%CI=1.54-1.67; Males IRR=1.51, 95%CI=1.47-1.56), in individuals with no TBI (IRR=1.54, 95%CI=1.50-1.59) or severe TBI (IRR=1.64, 95%CI=1.35-1.99), compared to mild TBI (IRR=1.36, 95%CI=1.26-1.48), and in children of fathers outside the workforce (IRR =2.44, 95%CI=1.75-3.39]; employed IRR=1.53, 95%CI=1.50-1.57). Further, the effect of ADHD-PRS on ADHD diagnosis was lower in children of mothers with an autoimmune disorder (IRR=1.19, 95%CI=1.05-1.35) than in mothers without an autoimmune disorder (IRR=1.53, 95%CI=1.49-1.57). This was also the only the interaction to survive false discovery rate (FDR) correction (*p*<0.009) for multiple testing (Table S3).

**Figure 2.**
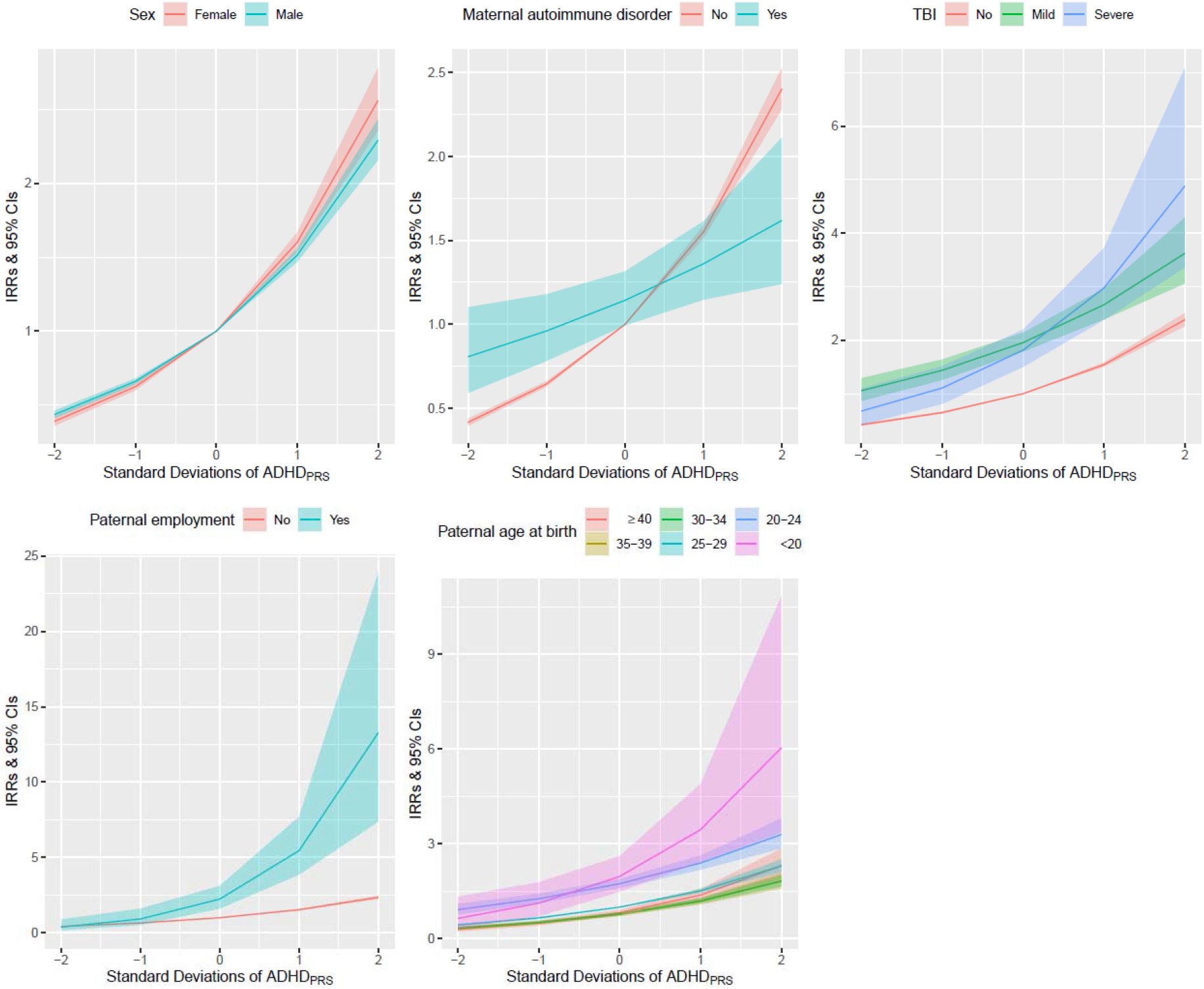
**Gene-environment interactions showing the differential linear effect of ADHD-PRS across levels of risk-factors on ADHD case-control status, expressed as incidence rate ratios (IRRs) with 95% confidence intervals (CIs)** **Note**: Incidence rate ratios (IRRs) and 95% confidence intervals (CIs) reflects the differential linear effect of a one standard deviation increase in ADHD-PRS across levels of each risk-factor, adjusted for sex, birth-year, and the first four PCs. Results are shown only for risk-factor showing tentative evidence of interaction with ADHD-PRS (p<0.05) in interaction analyses. Full results are presented in Table S3. For exposure definitions, see Table S1. **Abbreviations**: TBI, traumatic brain injury. PRS, polygenic risk score

## Discussion

We investigated three types of gene-environment interplay (association, confounding and interaction) in ADHD, using ADHD-PRS and a broad range of risk-factors. ADHD-PRS was associated with 14 out of 25 evaluated risk-factors, including certain birth-related and somatic risk-factors, and psychosocial risk-factors expect paternal employment in the general population. Nevertheless, ADHD-PRS and/or parental psychiatric history had minimal confounding effect on the phenotypic association of these risk-factors and ADHD diagnosis. Finally, we observed tentative support for gene-environment interactions for six risk-factors, including TBI, paternal employment and age at birth of index child.

We present a targeted review of prior literature linking ADHD to the risk-factors included in this study at the phenotypic, familial and common genetic variant level. We replicate majority of these associations in a population-based sample including 13,697 ADHD individuals. Moreover, we show that in the general population, higher ADHD-PRS is associated with exposure to several ADHD risk-factors, albeit with small effect sizes. Together with prior reserach,^11, 13^ this highlights that gene-environment associations appear to be pervasive in ADHD, particularly for psychosocial adversity. We replicate previous findings in iPSYCH2012^13^ of ADHD-PRS being associated with parental unemployment, lower education, income, and psychiatric disorders. We add to this by showing that higher ADHD-PRS is also associated with growing up in a single parent household and being born to young parents. Similar ADHD genetic correlations with lower educational attainment and younger age at child-birth have previously been reported in the UK Biobank (Table 1). Further, ADHD-PRS was associated with being small for gestational age, TBI, and infections in childhood, suggesting that these ADHD risk-factor association arise in part through shared genetic mechanisms.

Despite evidence for gene-environment associations, ADHD-PRS and parental history of psychiatric disorders had only minimal confounding effects on these associations between risk-factors and ADHD case-control status. However, we cannot rule out substantial residual genetic confounding as current ADHD-PRS only explains ∼5% of variance in ADHD^3^ and register coverage of parental ADHD diagnoses is limited as ADHD only recently stared being diagnosed in adults.^2, 4^ Our results highlight that adjusting for ADHD-PRS and parental psychiatric history is unlikely to fully capture familial/genetic confounding, suggesting that researchers aiming to identify causal risk-factors for ADHD should consider other genomic and/or family-based methods to address genetic confounding.^5, 28^ Further, our finding indicate that ADHD-PRS and parental psychiatric history act largely independently of the other investigated risk-factors in the development of ADHD.

We observed interactions between ADHD-PRS and sex, maternal autoimmune disease, TBI, and parental employment and age at child-birth. For instance, children with higher ADHD-PRS whose fathers were outside the workforce or young (<20 years) at child-birth had more elevated risk of ADHD, compared to those with none or only one of these risk-factors. These findings may in part be explained by undiagnosed ADHD in fathers, as younger parenthood and unemployment is linked to ADHD (Table 1). Unlike prior work in iPSYCH,^29^ we observed an interaction of ADHD-PRS with sex. However, the effect was small and should be interpreted with be given the large prevalence difference of ADHD in boys and girls.^2^ Overall, there was limited evidence of gene-environment interactions, in line with the few ADHD-PRS interaction studies conducted so far.^11^ It should also be noted that only the interaction of ADHD-PRS with maternal autoimmune disorder survived multiple-testing correction, and given that maternal autoimmune disorder did not have a main effect on ADHD, the results are difficult to interpret. Utilizing PRS to detect gene-environment interaction is challenging, both due to low power and because disorder predictive PRSs do not necessarily capture genetic variants linked to a differential susceptibility to risk exposure.^9^ Nevertheless, investigating gene-environment interplay may elucidate heterogeneous pathways to ADHD; we highlight three examples here. First, epilepsy was associated with a 2.3-fold increased risk of ADHD, but with no evidence for associations or interactions between epilepsy and ADHD-PRS. Prior studies also do not support a strong common genetic overlap between epilepsy and ADHD (Table 1), suggesting that epilepsy may act as an independent pathway to ADHD (e.g., through neuronal insults), or that the disorders associate due to common environmental and/or rare genetic risk-factors. Second, mild TBI has been linked to ADHD, yet it remains unclear whether the association is causal, as untreated ADHD is itself a risk for accidents and injuries.^30^ In support of this, we found higher ADHD-PRS to be associated with higher risk of TBI. We also observed that the effect of ADHD-PRS on ADHD risk was stronger in those with no or severe TBI. Similarly, one prior study found ADHD-PRS to be associated with higher ADHD traits in youth without TBI, but not in those with mild TBI.^31^ Together, this suggests that individuals with a mild TBI may require a lower genetic burden to be diagnosed with ADHD. Finally, our results add to a growing body of evidence linking ADHD to immune-related diseases (Table 1). ADHD-PRS was associated with maternal autoimmune disease and early life infections. Moreover, maternal autoimmune disease showed evidence of interaction with ADHD-PRS, potentially suggesting that autoimmune/inflammatory diseases impact on ADHD risk both through a shared genetic vulnerability and through neuro-inflammatory pathways.^32^

## Limitations

Our findings must be interpreted in light of several limitations. First, although this study represent the largest and most comprehensive investigation of gene-environment interplay using ADHD-PRS to date, we may still have been underpowered to detect interactions of modest effect sizes, and we were limited to studying risk factor recorded with sufficient coverage in the Danish national registers. Second, our sample included clinically treated ADHD individuals with an ICD-10 diagnosis F90.0, meaning our findings may not extend to the broader ADHD population. Third, our analyses were restricted to individuals of European ancestry, which may limit generalizability to populations of other ancestry. Third, studying interactions with PRS is challenging; if included SNPs do not interact with the exposures in a similar way, interactions may be diluted, and SNPs associated with ADHD case-control status are not necessarily the genetic variants most strongly linked to a potential differential susceptibility to risk-factor exposure.^9, 33^

## Conclusion

ADHD polygenic liability increases the risk of both ADHD diagnosis and exposure to risk-factors associated with ADHD. Our findings underscore the importance of accounting for genetic confounding in order to identify causal risk-factors, and highlight that ADHD-PRS and family psychiatric history are unlikely to fully capture such confounding. We present suggestive evidence for gene-environment interactions between ADHD-PRS and certain risk-factors, illustrating potential different pathways to ADHD. As genotyped samples with information on both ADHD and associated risk-factors increase, future research should explore the mechanism underlying observed gene-environment associations and interactions in ADHD using complimentary methods such as genome-wide by environment interaction studies or plasticity PRS.^9^

## Data Availability

Data is available on request and in accordance with Danish law.

## Acknowledgments

The iPSYCH team was supported by grants from the Lundbeck Foundation (R102-A9118, R155-2014-1724 and R248-2017-2003), the European Union’s FP7 Program (Grant No. 602805, “Aggressotype”), the European Union’s Horizon 2020 Program (Grant No. 667302, “CoCA”), NIMH (1U01MH109514-01 to Dr. Borglum), and the universities and university hospitals of Aarhus and Copenhagen. The Danish National Biobank resource was supported by the Novo Nordisk Foundation. High-performance computer capacity for handling and statistical analysis of iPSYCH data on the GenomeDK HPC facility was provided by the Center for Genomics and Personalized Medicine and the Centre for Integrative Sequencing, iSEQ, Aarhus University, Denmark (grant to ADB). Dr. Dalsgaard’s research is further supported by Helsefonden (grant no 19-8-0260) and the European Union’s Horizon 2020 Program (Grant No 847879). Dr. LaBianca acknowledges support from the Research Fund of the Mental Health Services – Capital Region of Denmark R4A92.

## Disclosures

Dr Ditte Demontis has received speaking fee from Takeda, outside the submitted work. Remaining authors reports no financial relationships with commercial interests.

## Notes

### Author Declarations

The iPSYCH2012 study was approved by the Danish Scientific Ethics Committee, the Danish Health Data Authority, the Danish Data Protection Agency, and Danish Newborn Screening Biobank Steering Committee. The Danish Scientific Ethics Committee, in accordance with Danish legislation, has, for this study, waived the need for informed consent in biomedical research based on existing biobanks.

