## Supplementary materials for "Interplay of ADHD polygenic liability with birth-related, somatic and psychosocial factors in ADHD - a nationwide study"

### Methods S1. Targeted literature review of ADHD associated birth-related, somatic and psychosocial risk-factors

To identify birth-related, somatic and psychosocial factors associated with ADHD and to put our research in context, we conducted a non-systematic, targeted review of the current phenotypic, familial and common genetic variant literature of birth-related, somatic and psychosocial factors linked to ADHD. We searched PubMed until May 2021 to identify the most recent meta-analyses, umbrella reviews, systematic reviews and consensus statements of ADHD risk-factors and correlates and snowballed from these. For phenotypic and familial associations, we preferentially present pooled (and when available, adjusted) risk-estimates from the largest available meta-analysis, followed by evidence from published systematic reviews. If such estimates were not available, we present estimates from large-scale population studies, prioritized based on sample size and the inclusion of clinically diagnosed cases. For gene-environment correlation and interactions, we present results of the largest available genetic correlation analyses based on linkage disequilibrium score regression (LDSC) (PMID: 25642630) and summarize the available ADHD-PRS literature, guided by a recently published systematic review (PMID: 33548493). We did not identify any genome-wide by environment interaction studies on ADHD to include in the overview. To limit the scope of our targeted literature review, we did not include twin studies or molecular genetic studies using other methods than LDSC or PRS.

Results of the literature review are presented in Table 1 in the main text.

### Table S1. International classification of disease (ICD) and Anatomical Therapeutic Chemical (ATC) codes and definitions of included ADHD risk-factors

| **Domain** | **Risk-factor** | **ICD-codes /ATC-codes** | **Definition** | **Reference** |
| --- | --- | --- | --- | --- |
| **Birth-related** | Sex | n/a | Male (ref) | n/a |
|  | Birth weight (BW) | n/a | Low BW <2500 g  Average BW 2500–3999 g (ref)  High BW ≥ 4000 g | - PMID: 25726514 |
|  | Small for gestational age (SGA) | n/a | SGA defined as being in the 10^th^ percentile of the birthweight distribution in the iPSYCH2012 subcohort, within each birth year and sex | PMID:  23026073 |
|  | Apgar score at 5 min | n/a | 10 (ref), < 10 | - PMID: 25726514 |
| **Somatic** | Maternal hypertension during pregnancy | **ICD-8:** 637.00, 637.03, 637.09,637.99, 637.04, 637.19, 762.19, 762.29, 762.39  **ICD-10:** O13, O14.0, O14.1, O14.2, O14.9, O16, O15.0–15.9 | ≥1 discharge diagnoses 20 weeks after gestation until birth of index child | PMID: 30785920 |
|  | Maternal infection during pregnancy | **ICD-8:** 038, 070, 000-009, 540, 680-686, 050-057, 110, 111, 035, 460-486, 580, 590, 59500, 59501, 612, 620, 622, 381-382, 04000-04399, 013, 320, 322, 392, 474, 04509-04699, 32300, 07199, 07202, 07501, 07929, 09049, 05201, 05302, 05403, 05501, 05601, 03609, 02701, 06209-06599, 09490-09499  **ICD-10:** A40, A41, B15-B19, A00-A09, K35, L00-L08, B00-B09, A46, J00-J18, N00, N10, N300, N390, N518B, N70, N71, N72, N76, N770D, N771B, N771L, H65-67, I02, G00-G07, A17, A80-89, B003, B004, B010, B011, B020, B021, B050, B051, B060, B261, B262, B375, B451, B582, B602, A022C, A548A, A548D, A521A, A521B, A229C, A321, A504, A390, E236A | ≥1 discharge diagnoses within 40 weeks prior to birth of index child | PMID: 30446204 |
|  | Maternal and paternal autoimmune disease | **ICD-8:** 242, 245.03, 249, 255.10, 255.11, 255.12, 255.18, 255.19,269, 281.00, 281.01, 281.08, 281.09, 283.90, 283.91, 446.49, 340, 354, 364, 446.29, 563.01, 563.19, 571.9, 571.93, 694, 696.09, 696.10, 696.19, 704, 709.01, 712.19, 712.39, 712.59 712.09, 716, 446.30, 446.31, 446.39, 733.09, 734.00, 734.01, 734.02, 734.08, 734.09, 734.19, 734.9, 712.49  **ICD-10:** E05.0, E06.3, E10, E27.1, K90.0, D51.0, D59.1, D69.3, G35, G61.0, H20, M31.3, K50, K51, K74.3, K73, L12, L10, L40 (except L40.4), L63, L80.9, M05, M06, M08, M33, M31.5, M31.6, M35.3, G70.0, M34, M32.1, M32.9, M35.0, M45.9 | ≥1 discharge diagnoses by child’s 5^th^ birthday | - PMID: 28219489 |
|  | Infections | See maternal infection during pregnancy | Time-varying, first and 5^th^ discharge diagnoses | - PMID: 30446204 |
|  | Asthma | **ICD-8:** 493  **ICD‐10:** J45.0, J45.1, J45.8, J45.9, J46.0, J46.9  **ATC:**  R03BA01-R03BA08, R03DC01-R03DC04, R03DC03, R03DB04, R03DA54, R03BB01, R03DX05 | Time-varying, ≥1 discharge diagnosis and/or ≥ 1 prescriptions filled within a year | - PMID: - 25828267   PMID: 28632331 |
|  | Atopic disease (atopic dermatitis, allergic rhinitis) | **ICD-8:** 691, 507, 502.00  **ICD-10:** L20, L308C, J30, J31.0  **ATC:** ≥1 prescription of D11AH, S01GX, V01A  ≥2 prescriptions of D07, R01AD01 – R01AD60, R06A | Time-varying, first discharge diagnosis | - PMID: - 25828267   PMID: 28632331 |
|  | Type 1 diabetes | **ICD-8:** 250.00–250.09 diagnosed before age 18  **ICD-10:** E10 diagnosed before age 18 | Time-varying, first discharge diagnosis | - PMID: - 33824142 |
|  | Epilepsy | **ICD-8:** 345 (excluding 345.29)  **ICD-10:** G40 | Time-varying, first discharge diagnosis | - PMID: 27412639 |
|  | Traumatic brain injury (TBI) | **ICD-8:** 850, **ICD-10,** S06.0 (**Mild TBI**)  **ICD-8:** 800-804, 851-854 **(Moderate/severe TBI**)  **ICD-10:** S02.0-1, S02.3, S02.7-9, S06.1-9, S07.0-S07.1, S07.8-S07.9, S09.0, S09.7-9, S18, T02.0, T04.0, T06.0, T90.2, T90.5, T90.8-9 **(Moderate/severe TBI**) | Time-varying, first discharge diagnosis | - PMID: 28301451 - PMID: 27552147 |
| **Psychosocial** | Maternal and paternal income | n/a | Defined by quintiles in the subcohort at index child’s year of birth | - PMID: 26249301 |
|  | Maternal and paternal education | n/a | Highest completed level of education at index child’s year of birth (elementary school, high school, or academic degree) | - PMID: 26249301 |
|  | Maternal and paternal employment | n/a | Defined as outside of the workforce, including retirement and currently in education which are both uncommon in new parents (ref), or employed in index child’s birth year | - PMID: 26249301 |
|  | Living in a single parent household | n/a | Evaluated in the first five years of life (no years=ref) | - PMID: 27355346 |
|  | Maternal and paternal age at birth | n/a | Maternal age: <20, 20-24, 25-29 (ref), 30-34, ≥35 years-of-age  Paternal age: <20, 20-24, 25-29, 30-34 (ref), 35-39, ≥40 years-of-age | - PMID: 24452535 |
|  | Parental psychiatric history | **ICD-8:** 290-315  **ICD-10:** F00-F99 | ≥1 discharge diagnoses for any psychiatric disorder in mother, father, or both by index child’s 5^th^ birthday | - PMID: 27355346 |

**Note:** Diagnoses in the Danish National Patient Register (DNPR) and the Danish Psychiatric Central Research Register (DPCRR) are recorded according to the International classification of disease (ICD) version 8 between 1977 and 1993 and ICD-10 from 1994 onwards. ICD-9 was never implemented in Denmark. The Danish National Prescription Registry (DPR) includes information on all prescriptions redeemed at Danish pharmacies since 1995 coded according to the Anatomical Therapeutic Chemical (ATC) classification system.

**Abbreviations:** Na, not applicable. Ref, reference category in statistical analyses.

### Table S2. Associations of ADHD-PRS with evaluated risk-factors in the iPSYCH2012 subcohort (N=21,578), expressed as odds ratios (OR) with 95% confidence intervals (CIs)

| Domain | Risk factor | Level | N | OR per PRS SD | Lower CI | Upper CI |
| --- | --- | --- | --- | --- | --- | --- |
| Birth-related | Sex | Male | 10994 | 1 (ref) |  |  |
|  |  | Female | 10584 | 0.98 | 0.95 | 1.00 |
|  | Birth weight | <2500 g | 698 | 1.02 | 0.95 | 1.10 |
|  |  | 2500-3999 g | 16865 | 1 (ref) |  |  |
|  |  | ≥ 4000 g | 3932 | 0.99 | 0.95 | 1.02 |
|  | Small for gestational age | No | 19564 | 1 (ref) |  |  |
|  |  | Yes | 1931 | **1.08** | **1.03** | **1.13** |
|  | Apgar score at 5 minutes | 10 | 19892 | 1 (ref) |  |  |
|  |  | <10 | 1487 | 1.01 | 0.95 | 1.06 |
| Somatic | Maternal hypertensive disorders during pregnancy | No | 21264 | 1 (ref) |  |  |
|  |  | Yes | 314 | 1.01 | 0.90 | 1.12 |
|  | Maternal infection during pregnancy | No | 20952 | 1 (ref) |  |  |
|  |  | Yes | 626 | 1.06 | 0.98 | 1.15 |
|  | Maternal autoimmune disorder by child’s 5^th^ birthday | No | 21012 | 1 (ref) |  |  |
|  |  | Yes | 566 | **1.14** | **1.04** | **1.24** |
|  | Paternal autoimmune disorder by child’s 5^th^ birthday | No | 21193 | 1 (ref) |  |  |
|  |  | Yes | 385 | 0.98 | 0.88 | 1.08 |
|  | 1 infection | No | 13245 | 1 (ref) |  |  |
|  |  | Yes | 8333 | **1.07** | **1.05** | **1.10** |
|  | ≥ 5 infections | No | 20989 | 1 (ref) |  |  |
|  |  | Yes | 589 | **1.14** | **1.05** | **1.24** |
|  | Asthma | No | 18688 | 1 (ref) |  |  |
|  |  | Yes | 2890 | **1.04** | **1.00** | **1.08** |
|  | Atopic diseases | No | 19814 | 1 (ref) |  |  |
|  |  | Yes | 1764 | 0.97 | 0.93 | 1.02 |
|  | Type 1 diabetes | No | 21486 | 1 (ref) |  |  |
|  |  | Yes | 92 | 0.89 | 0.72 | 1.09 |
|  | Epilepsy | No | 21137 | 1 (ref) |  |  |
|  |  | Yes | 441 | 0.99 | 0.90 | 1.09 |
|  | Traumatic brain injury | No | 19780 | 1 (ref) |  |  |
|  |  | Mild | 1558 | **1.11** | **1.05** | **1.17** |
|  |  | Severe | 339 | 1.09 | 0.98 | 1.21 |
| Psychosocial | Maternal income at birth | 1. quintile | 4302 | **1.19** | **1.14** | **1.24** |
|  |  | 2. quintile | 4318 | **1.19** | **1.14** | **1.24** |
|  |  | 3. quintile | 4315 | **1.10** | **1.05** | **1.15** |
|  |  | 4. quintile | 4317 | **1.04** | **1.00** | **1.09** |
|  |  | 5. quintile | 4326 | 1 (ref) |  |  |
|  | Paternal income at birth | 1. quintile | 4277 | **1.19** | **1.14** | **1.24** |
|  |  | 2. quintile | 4293 | **1.14** | **1.10** | **1.19** |
|  |  | 3. quintile | 4294 | **1.09** | **1.04** | **1.13** |
|  |  | 4. quintile | 4293 | **1.04** | **1.00** | **1.09** |
|  |  | 5. quintile | 4302 | 1 (ref) |  |  |
|  | Maternal education at birth | Elementary | 6022 | **1.16** | **1.12** | **1.20** |
|  |  | High school | 9409 | 1 (ref) |  |  |
|  |  | Academic degree | 5914 | **0.89** | **0.86** | **0.92** |
|  | Paternal education at birth | Elementary | 5001 | **1.17** | **1.13** | **1.21** |
|  |  | High school | 10926 | 1 (ref) |  |  |
|  |  | Academic degree | 5002 | **0.89** | **0.86** | **0.92** |
|  | Maternal employment at birth | In | 21472 | 1 (ref) |  |  |
|  |  | Out of | 99 | **1.22** | **1.01** | **1.49** |
|  | Paternal employment at birth | In | 21313 | 1 (ref) |  |  |
|  |  | Out of | 97 | 1.11 | 0.91 | 1.36 |
|  | Living in a single parent household by child’s 5^th^ birthday | No | 17384 | 1 (ref) |  |  |
|  |  | 1 year | 1240 | **1.14** | **1.07** | **1.20** |
|  |  | 2 years | 895 | **1.21** | **1.13** | **1.30** |
|  |  | 3 years | 793 | **1.28** | **1.19** | **1.38** |
|  |  | 4 years | 616 | **1.30** | **1.20** | **1.41** |
|  |  | 5 years | 650 | **1.26** | **1.16** | **1.36** |
|  | Maternal age at birth | <20 | 412 | **1.29** | **1.17** | **1.42** |
|  |  | 20-24 | 3912 | **1.15** | **1.11** | **1.20** |
|  |  | 25-29 | 8543 | 1 (ref) |  |  |
|  |  | 30-34 | 6283 | 0.97 | 0.94 | 1.01 |
|  |  | ≥35 | 2410 | **0.95** | **0.90** | **0.99** |
|  | Paternal age at birth | <20 | 112 | **1.25** | **1.04** | **1.50** |
|  |  | 20-24 | 1972 | **1.22** | **1.16** | **1.28** |
|  |  | 25-29 | 6880 | **1.04** | **1.01** | **1.08** |
|  |  | 30-34 | 7380 | 1 (ref) |  |  |
|  |  | 35-39 | 3630 | 1.00 | 0.96 | 1.04 |
|  |  | ≥40 | 1511 | 0.96 | 0.91 | 1.01 |
|  | Parental history of psychiatric disorder by child’s 5^th^ birthday | No | 20281 | 1 (ref) |  |  |
|  |  | One parent | 1211 | **1.13** | **1.07** | **1.20** |
|  |  | Both parents | 86 | **1.30** | **1.05** | **1.61** |

**Note**: Odds ratios and 95% CI reflect the increase in risk of exposure by one per standard deviation increase in the ADHD-PRS. Significant associations are highlighted in bold.

**Abbreviations**: OR, odds ratio. LCI, lower 95% confidence interval. UCI, upper 95% confidence interval.

### Table S3. Interaction between each putative risk-factor and ADHD-PRS on ADHD case-controls status, showing the differential (linear) effect of PRS per SD across levels of risk factors for ADHD

| Domain | Risk factor | Level | IRR | LCI | LCI | Interaction p-value | FDR  p-value |
| --- | --- | --- | --- | --- | --- | --- | --- |
| Birth-related | Sex | Female | 1.60 | 1.54 | 1.67 |  |  |
|  |  | Male | 1.51 | 1.47 | 1.56 | **0.03** | 0.67 |
|  | Birth weight | <2500 g | 1.77 | 1.55 | 2.03 |  |  |
|  |  | 2500-3999 g | 1.54 | 1.49 | 1.58 |  |  |
|  |  | ≥ 4000 g | 1.49 | 1.41 | 1.58 | 0.07 | 0.67 |
|  | Small for gestational age | No | 1.54 | 1.50 | 1.58 |  |  |
|  |  | Yes | 1.53 | 1.41 | 1.65 | 0.86 | 1 |
|  | Apgar score at 5 minutes | 10 | 1.54 | 1.50 | 1.58 |  |  |
|  |  | <10 | 1.46 | 1.34 | 1.59 | 0.24 | 1 |
| Somatic | Maternal hypertensive disorders during pregnancy | No | 1.54 | 1.50 | 1.57 |  |  |
|  |  | Yes | 1.60 | 1.28 | 1.98 | 0.73 | 1 |
|  | Maternal infection during pregnancy | No | 1.53 | 1.50 | 1.57 |  |  |
|  |  | Yes | 1.50 | 1.32 | 1.70 | 0.71 | 1 |
|  | Maternal autoimmune disorder by child’s 5^th^ birthday | No | 1.55 | 1.51 | 1.59 |  |  |
|  |  | Yes | 1.19 | 1.05 | 1.35 | **<.0001** | **<.01** |
|  | Paternal autoimmune disorder by child’s 5^th^ birthday | No | 1.53 | 1.49 | 1.57 |  |  |
|  |  | Yes | 1.77 | 1.47 | 2.15 | 0.13 | 1 |
|  | 1 infection | No | 1.55 | 1.50 | 1.60 |  |  |
|  |  | Yes | 1.49 | 1.43 | 1.55 | 0.12 | 1 |
|  | ≥ 5 infections | No | 1.53 | 1.50 | 1.57 |  |  |
|  |  | Yes | 1.49 | 1.30 | 1.70 | 0.66 | 1 |
|  | Asthma | No | 1.54 | 1.50 | 1.58 |  |  |
|  |  | Yes | 1.48 | 1.39 | 1.58 | 0.24 | 1 |
|  | Atopic diseases | No | 1.54 | 1.50 | 1.58 |  |  |
|  |  | Yes | 1.51 | 1.30 | 1.65 | 0.72 | 1 |
|  | Type 1 diabetes | No | 1.53 | 1.50 | 1.57 |  |  |
|  |  | Yes | 2.08 | 1.42 | 3.06 | 0.12 | 1 |
|  | Epilepsy | No | 1.54 | 1.50 | 1.58 |  |  |
|  |  | Yes | 1.42 | 1.22 | 1.66 | 0.32 | 1 |
|  | Traumatic brain injury | No | 1.54 | 1.50 | 1.59 |  |  |
|  |  | Mild | 1.36 | 1.26 | 1.48 |  |  |
|  |  | Severe | 1.64 | 1.35 | 1.99 | **0.01** | 0.37 |
| Psychosocial | Maternal income at birth | 1. quintile | 1.48 | 1.41 | 1.56 |  |  |
|  |  | 2. quintile | 1.48 | 1.40 | 1.55 |  |  |
|  |  | 3. quintile | 1.54 | 1.45 | 1.63 |  |  |
|  |  | 4. quintile | 1.57 | 1.48 | 1.67 |  |  |
|  |  | 5. quintile | 1.49 | 1.39 | 1.59 | 0.50 | 1 |
|  | Paternal income at birth | 1. quintile | 1.52 | 1.45 | 1.60 |  |  |
|  |  | 2. quintile | 1.42 | 1.35 | 1.50 |  |  |
|  |  | 3. quintile | 1.50 | 1.42 | 1.59 |  |  |
|  |  | 4. quintile | 1.59 | 1.50 | 1.68 |  |  |
|  |  | 5. quintile | 1.56 | 1.46 | 1.67 | 0.07 | 0.67 |
|  | Maternal education at birth | Elementary | 1.46 | 1.40 | 1.52 |  |  |
|  |  | High school | 1.53 | 1.47 | 1.59 |  |  |
|  |  | Academic degree | 1.41 | 1.34 | 1.49 | 0.06 | 0.67 |
|  | Paternal education at birth | Elementary | 1.46 | 1.40 | 1.53 |  |  |
|  |  | High school | 1.50 | 1.45 | 1.55 |  |  |
|  |  | Academic degree | 1.48 | 1.38 | 1.58 | 0.67 | 1 |
|  | Maternal employment at birth | In | 1.54 | 1.50 | 1.58 |  |  |
|  |  | Out of | 1.32 | 0.95 | 1.85 | 0.38 | 1 |
|  | Paternal employment at birth | In | 1.53 | 1.50 | 1.57 |  |  |
|  |  | Out of | 2.44 | 1.75 | 3.39 | **0.006** | 0.28 |
|  | Living in a single parent household the first five years of life | No | 1.53 | 1.48 | 1.58 |  |  |
|  |  | 1 year | 1.45 | 1.32 | 1.59 |  |  |
|  |  | 2 years | 1.40 | 1.26 | 1.56 |  |  |
|  |  | 3 years | 1.41 | 1.26 | 1.57 |  |  |
|  |  | 4 years | 1.35 | 1.21 | 1.51 |  |  |
|  |  | 5 years | 1.37 | 1.22 | 1.54 | 0.06 | 0.67 |
|  | Maternal age at birth | <20 | 1.54 | 1.32 | 1.79 |  |  |
|  |  | 20-24 | 1.43 | 1.35 | 1.51 |  |  |
|  |  | 25-29 | 1.53 | 1.47 | 1.59 |  |  |
|  |  | 30-34 | 1.50 | 1.43 | 1.58 |  |  |
|  |  | ≥35 | 1.60 | 1.48 | 1.74 | 0.15 | 1 |
|  | Paternal age at birth | <20 | 1.75 | 1.30 | 2.36 |  |  |
|  |  | 20-24 | 1.38 | 1.28 | 1.48 |  |  |
|  |  | 25-29 | 1.52 | 1.45 | 1.59 |  |  |
|  |  | 30-34 | 1.53 | 1.46 | 1.60 |  |  |
|  |  | 35-39 | 1.54 | 1.44 | 1.64 |  |  |
|  |  | ≥40 | 1.67 | 1.51 | 1.86 | **0.04** | **0.67** |
|  | Parental history of psychiatric disorder by child’s 5^th^ birthday | No | 1.54 | 1.50 | 1.58 |  |  |
|  |  | One parent | 1.44 | 1.31 | 1.57 |  |  |
|  |  | Both parents | 1.74 | 1.18 | 2.57 | 0.30 | 1 |

Note: Significant interactions are highlighted in bold.

Abbreviations: IRR, incidence rate ratio. LCI, lower 95% confidence interval. UCI, upper 95% confidence interval.

### Figure S1. Flowchart of study population selection

**iPSYCH2012 population**

18 726 ADHD individuals

29 613 subcohort

**Subject level genotype exclusion criteria** (N excluded 12 475)

Missing genotypes/failed SNP QC

Non-European ancestry

Related individuals closer than 3^rd^ degree

**Inclusion criteria** (N excluded 877)

Alive and living in Denmark by age 5

**Eligible study population**

17 745 ADHD individuals outside subcohort

374 ADHD individuals in subcohort

29 343 non-ADHD individuals in subcohort

**Final study population for analyses**

13 409 ADHD individuals outside subcohort

288 ADHD individuals in subcohort

21 290 non-ADHD individuals in subcohort

### Figure S2. Associations of ADHD-PRS (A) and parental psychiatric history (B) with ADHD case-control status, expressed as incidence rate ratios (IRRs) with 95% confidence intervals (CIs)

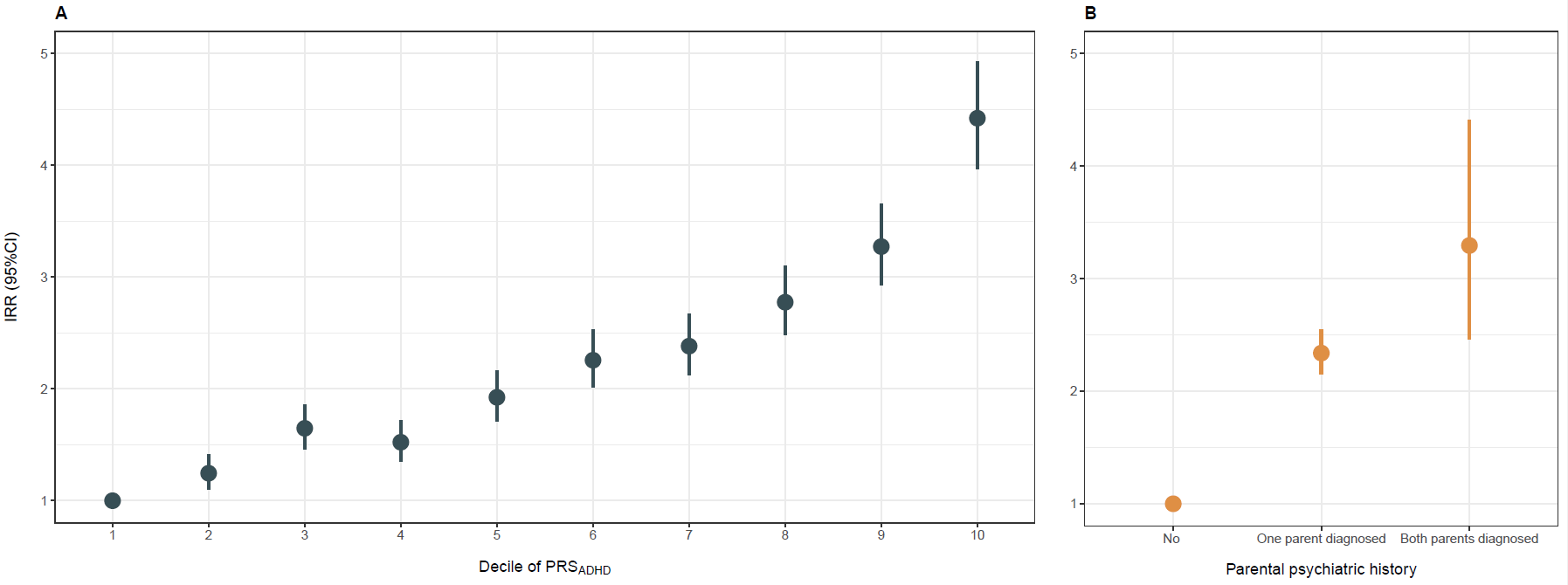

Note: Incidence ratio ratios and 95%CI reflect the increase in risk of ADHD by A) deciles of the ADHD-PRS, and B) parental history of any clinically diagnosed psychiatric disorder.

Abbreviations: IRR, incidence rate ratio. LCI, lower 95% confidence interval. UCI, upper 95% confidence interval. PRS, polygenic risk score.
